# Uptake of intramuscular vitamin K administration after birth and maternal and infant demographic variables: a national cohort study

**DOI:** 10.1101/2023.02.27.23286516

**Authors:** S Brunton, L Fenton, P Hardelid, TC Williams

## Abstract

A long-acting monoclonal antibody against Respiratory Syncytial Virus (RSV), given as a one-off injection shortly after birth, is likely to be introduced soon. We hypothesised that carer acceptance of intra-muscular (IM) vitamin K, another injection given shortly after birth, might serve as a proxy indicator of likely acceptance of any such anti-RSV intervention, given previous associations described between IM vitamin K acceptance and subsequent non-immunisation. Using a national dataset of all postnatal health visitor visits in Scotland from 2018-2021 we explored demographic variables associated with non-acceptance of IM vitamin after birth. We found that in the time period 2019-2021 over 95.5% of carers were documented as consenting to this intervention, with only 1.1% requesting oral vitamin K and 0.9% refusing vitamin K altogether. Infant ethnicity, use of English as a first language at home, socio-economic position and maternal age were not associated with reduced uptake of IM vitamin K. We therefore did not identify any groups that might require increased engagement prior to the roll-out of a long-acting monoclonal antibody for RSV.

## Introduction

Following recent positive results for nirsevimab, a long-acting monoclonal against RSV, in a phase 3 trial^1^, it is likely to be shortly introduced to the infant population in Europe and the United States. As a one-off injection of an intra-muscular (IM) agent potentially given shortly after birth, it bears more similarity to the routine administration of Vitamin K than to those of routine infant immunisations, which in the United Kingdom commence at the age of 2 months^2^.

In the United Kingdom, vitamin K can be administrated as an IM (routine administration) or an oral agent (which must be prescribed by a medical professional). A review found that carers who do not consent to the administration of IM vitamin K, request oral vitamin K instead, or refuse all vitamin K, may do so for two broad reasons^3^. The first is due to concerns about the potential harm to an infant: the distress that injection for their newborn child may cause, or because of concerns that IM vitamin K may lead to an increased risk of leukaemia, a possibility raised in the 1990s^4^ that was subsequently refuted by further research^5,6^. A second category of carers may desire their infants to have a more “natural” neonatal course, driven either by religious beliefs or a belief that vitamin K deficiency is to be expected for infants^3^.

Previous studies have found conflicting relationships between carer demographics and refusal of IM vitamin K. A study from the United States showed that mothers who refused IM vitamin K were more likely to be of White ethnicity, aged >30 years, or college graduates^7^; however other studies found maternal non-white ethnicity to be a predictor for refusal of vitamin K^8,9^.

Decisions made about modality of vitamin K administration appear to be associated with subsequent choices about childhood immunisations. A study from the United States found an association between the refusal of IM vitamin K and the refusal of other preventive measures, including the hepatitis B vaccine at birth, prophylaxis against gonococcal ophthalmia, and subsequent routine vaccination^10^. A study from New Zealand found that carers who declined vitamin K for their newborn infants were 14-fold more likely to not proceed with routine childhood immunisations, and those that opted for oral vitamin K were 3.5 times more likely to subsequently non-immunise^8^.

With this study we aimed to understand the uptake of IM vitamin K in infants in Scotland. We examined the association between maternal and infant demographic variables and uptake to understand whether there are populations for which the use of a novel IM agent shortly after birth may be less acceptable. We hypothesised that this information, as well as being informative about the overall acceptability of an intervention such as nirsevimab, could inform decisions on which populations might need specific targeted interventions to boost the uptake of an immunisation such as nirsevimab.

## Materials and methods

### Dataset

All variables described below are collected as part of the Health Visitor First Visit Report, which is offered at the age of 10-14 days for all newborn infants in Scotland. Coverage of the health visitor first visit is high, with 97.2% of eligible children (those turning 10 days old) receiving a review in the period 2018-2021^11^. Data collected includes date of birth, address with postcode, gender, maternal age, and the first language used at home (Supplementary File 1). Data was analysed for infants born between 1 January 2018 to 31 December 2022. Specific ethical approval was not considered necessary as this study made use of existing aggregate population-level data, without any linkage or access to individual records.

### Variables collected

We collected information on the primary outcome measures of IM vitamin K administration, oral vitamin K administration, or no Vitamin K administration.

We also collected information on explanatory variables of maternal age (age bands <20, 20-24, 25-29, 30-34, 35-40 and >40), infant ethnicity (white Scottish, white other British, white Polish, white Other, Asian, Black Caribbean/African, Mixed/Multiple Ethnicity, and Other/Not Known), whether English was a first language at home (Yes/ No) and Scottish Index of Multiple Deprivation^12^, a relative measure of deprivation across 6,976 small areas in Scotland called data zones (with a mean of 785 individuals per data zone in 2021), based on home postcode, grouped by quintile.

### Analysis

We first explored patterns of missing recording of Vitamin K administration. We tested to see whether there were differences over time in the proportion of infants who received IM vitamin K, as there may have been behavioural differences before and after the Covid-19 pandemic. We used a Fisher’s exact test to compare the proportion of infants who received IM vitamin K in 2018 and 2019 to subsequent years (2019-2021 and 2020-2021 respectively). We grouped together oral and no vitamin K administration as markers of IM vitamin K non-acceptance and compared this to IM vitamin K administration.

We then examined whether there were associations between IM vitamin K administration or oral/no vitamin K and each explanatory variable. We used a Fisher exact test for categorical variables that could be set out in a 2 × 2 contingency table (English as a first language at home), and Chi square testing for categorical variables with a larger number of variables (ethnicity, SIMD and maternal age). All analyses were implemented in RStudio and R version 4.2.2^13^. Other/Not Known ethnicity was excluded as this group was likely to be highly heterogenous. Missing/incomplete data was not included in the analyses, and no imputation was made of missing data as this was unlikely to be missing randomly. All scripts and data produced are available online at https://git.ecdf.ed.ac.uk/twillia2/vitk_administration_scotland/

### Ethics

This analysis was undertaken on aggregate routinely-collected data obtained from Public Health Scotland via information request and released in accordance with PHS disclosure control procedures. More information on how PHS collects and processes health data is available via their privacy notice^14^.

### Funding

No specific funding was received for this project; TCW is funded by the Wellcome Trust. Research at UCL Great Ormond Street Institute of Child Health is supported by the NIHR GOSH Biomedical Research Centre.

## Results

### Features of the dataset

Data were available for a total of 193,441 infants reviewed from 2018 to 2021 (Table 1). The percentage of newborns who received IM vitamin K increased from 93.7% in 2018 to 95.7% in 2021. The likelihood of IM vitamin K being administered increased from 2018 compared to subsequent years (2019-2021, OR 1.72, p< 0.001) but there was no further significant increase from 2019 onwards (OR of 1.03 for 2019 compared to 2020-2021, p=0.48). Therefore subsequent analyses were conducted using the dataset from 2019 to 2021 (142,857 infants). This 2019-2021 dataset had low rates of missing or invalid data (2.7%) and overall 1.8% of carers opted for either oral vitamin K (1.1%) or no vitamin K (0.7%).

**Table 1.** Vitamin K administration data for the period 2018 to 2021.

| Vit K administration | 2018 (%) | 2019 (%) | 2020 (%) | 2021 (%) | Totals for 2019-2021 (%) |
| --- | --- | --- | --- | --- | --- |
| Intramuscular | 47,419 (93.7) | 46,644 (95.1) | 44,886 (95.7) | 44,891 (95.7) | 136,421 (95.5) |
| Oral | 1,086 (2.1) | 569 (1.2) | 542 (1.2) | 514 (1.1) | 1,625 (1.1) |
| None | 455 (0.9) | 330 (0.7) | 304 (0.6) | 319 (0.7) | 953 (0.7) |
| Missing/invalid | 1,624 (3.2) | 1,527 (3.1) | 1,166 (2.5) | 1,165 (2.5) | 3,858 (2.7) |
| **Total** | **50,584** | **49,070** | **46,898** | **46,889** | **142,857** |

**Table 2:**
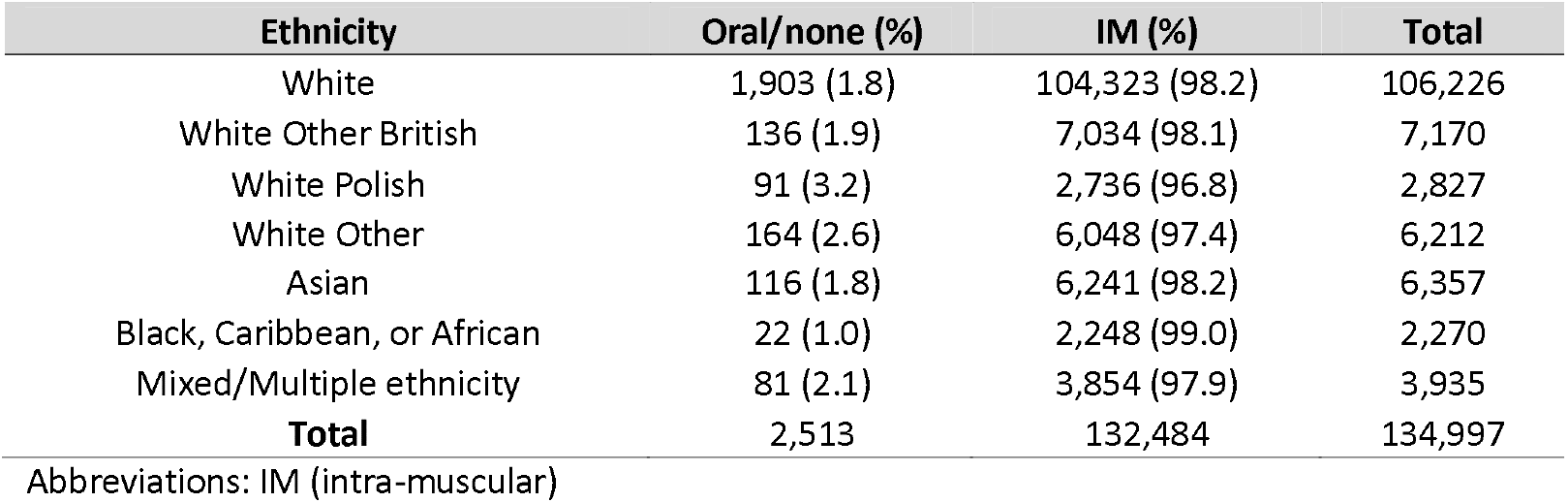
Vitamin K administration data for the period 2019-2021 by infant ethnicity.

| Ethnicity | Oral/none (%) | IM (%) | Total |
| --- | --- | --- | --- |
| White | 1,903 (1.8) | 104,323 (98.2) | 106,226 |
| White Other British | 136 (1.9) | 7,034 (98.1) | 7,170 |
| White Polish | 91 (3.2) | 2,736 (96.8) | 2,827 |
| White Other | 164 (2.6) | 6,048 (97.4) | 6,212 |
| Asian | 116 (1.8) | 6,241 (98.2) | 6,357 |
| Black, Caribbean, or African | 22 (1.0) | 2,248 (99.0) | 2,270 |
| Mixed/Multiple ethnicity | 81 (2.1) | 3,854 (97.9) | 3,935 |
| **Total** | **2,513** | **132,484** | **134,997** |

Abbreviations: IM (intra-muscular)

### Infant ethnicity and English as a first language in the home

Full data was available on ethnicity for 134,997 infants (94.5%) from 2019 to 2021. IM vitamin K non-acceptance was highest in the white Polish group (3.2% of those with data available opted for oral or no vitamin K) and lowest in the Black, African or Caribbean group (1.0% opted for oral or no vitamin K); however sample sizes were small both groups (2,827 and 2,270 infants respectively) and a chi-square test showed no evidence of a significant difference between the groups (p=0.23).

Data was available on the first language used at home for 133,599 infants (93.5%). For those with English as a first language 1.8% opted for oral or no vitamin K, as opposed to 2.1% for those who did not state that English was their first language; this difference was not significant (p=0.06).

### Area-level deprivation

Data was available on socioeconomic position categorised by SIMD quintile for 138,958 infants (97.2%). There was no clear association between socio-economic position and IM vitamin K acceptance (Table 3; p=0.22).

**Table 3.** Scottish Index of Multiple Deprivation status and vitamin K administration for the period 2019-2021.

| SIMD Quintile | Oral/none (%) | IM (%) |
| --- | --- | --- |
| Q1 (most deprived) | 556 (1.7) | 32,101 (98.3) |
| Q2 | 650 (2.3) | 28,230 (97.7) |
| Q3 | 596 (2.3) | 24,806 (97.7) |
| Q4 | 468 (1.6) | 27,911 (98.4) |
| Q5 (least deprived) | 308 (1.3) | 23,332 (98.7) |
| **Total** | **2,578** | **136,380** |

Abbreviations: SIMD (Scottish Index of Multiple Deprivation); IM (intra-muscular)

### Maternal age

Data was available on maternal age for 138,868 infants (97.2%). The maternal age bands with the highest rates of IM vitamin K acceptance were 30-34 and 35-39. However absolute differences between the groups with the highest (<20 years; 2.3%) and lowest (30-34 years; 1.6%) rates of IM vitamin K non-acceptance was small (0.7%), and there was no significant difference between the age distributions of those who opted for oral or no vitamin K, and those who opted for IM administration (p=0.22).

**Table 4.** Maternal age and vitamin K administration for the period 2019-2021.

| Maternal age | Oral/none (%) | IM (%) |
| --- | --- | --- |
| <20 | 79 (2.3) | 3,404 (97.7) |
| 20-24 | 382 (2.2) | 17,258 (97.8) |
| 25-29 | 769 (2.1) | 36,665 (97.9) |
| 30-34 | 745 (1.6) | 46,330 (98.4) |
| 35-39 | 474 (1.7) | 26,698 (98.3) |
| 40+ | 124 (2.0) | 5,940 (98.0) |
| **Total** | **2,573** | **136,295** |

Abbreviations: IM (intra-muscular)

## Discussion

Overall rates of IM vitamin K acceptance in this cohort were high, with only 1.8% of carers in this dataset opting for the oral administration of vitamin K (1.1%), or no vitamin K (0.7%). We did not find an association between baby ethnicity, English as a first language, socio-economic position as defined by SIMD quintile, maternal age, and IM vitamin K non-acceptance.

Our estimate is of 1.8% of carers in this dataset opting out of the recommended post-natal administration of IM vitamin K. These rates are higher than those from previous population-level studies in a high-income setting. A previous study analysed births in the province of Alberta in Canada in the period 2006-2012, where 0.72% of carers declined IM vitamin K for their newborn infant^15^, and another that examined IM vitamin K administration across the United States in 2015 similarly found that 0.6% of carers of refused this^9^. However, rates of vitamin K administration appear to be much lower in lower-income settings such India, where 62.4% of newborns received this postnatally^16^, or Nepal, where only 17.1% of infants in government hospitals received vitamin K^17^, and therefore our findings are unlikely to be applicable to such settings, where systems and access factors are likely to have an influence as well as acceptability.

Strengths of our study include population level data for postnatal visits for infants born in all settings, very low rates of missing data, and up-to-date information on this intervention in a post-pandemic setting. The Scottish Child Health Surveillance Programme is unique in the United Kingdom for its coverage and completeness and therefore our findings are likely to be highly representative of the demographics of the births in this cohort during this time period.

Limitations include that this study was based on postnatal, aggregate data provided from the Child Health Surveillance Programme database by Public Health Scotland. Not all visits scheduled in the period 0-10 days take place, although in the period 2018-2021 97.3% of these did; infants likely not be included are those who are still in hospital at 10 days of age, or those whom the Health Visitor was unable to contact; IM vitamin K acceptance patterns in these groups may differ to those of the population as a whole.

In addition vitamin K IM acceptance profiles were not categorised according to birth location (home/birthing centre/hospital. A previous study from Tennessee in the United States showed an association between delivery at home or at a midwife-led birthing centre and reduced rates of IM vitamin K acceptance, with up to 14.5% of carers opting out of IM vitamin K at home deliveries and 31% in birthing centres^18^. Birth centres and maternity wards in Scotland are commonly co-located on the same hospital site, and births coded by site rather than exact location of delivery, so this data was not available for this analysis. Home births are recorded in a separate birth dataset (National Records of Scotland) to that used for this study so again this data was not available for this analysis.

Another limitation is that a high proportion of the mothers in this dataset self-reported white ethnicity (90.7%) and therefore these findings may not be applicable to other more ethnically diverse settings. Immunisation behaviours are known to differ between Scotland and adjacent England^19^, and therefore our findings may not necessarily be representative of the United Kingdom as a whole, or other high income settings.

## Data Availability

All data produced are available online at https://git.ecdf.ed.ac.uk/twillia2/vitk_administration_scotland/

## Conclusion

Rates of carer acceptance of an intramuscular injection of IM vitamin K after birth were very high in this national cohort. Infant ethnicity, use of English as a first language at home, socio-economic status and maternal age were not associated with reduced uptake of IM vitamin K. If vitamin K is a valid proxy marker for acceptance of a long-acting monoclonal anti-RSV antibody, then we did not identify any groups that might require increased engagement prior to the roll-out of such an intervention.

## Competing interests

Thomas C Williams is Principal Investigator for the BronchStart project, which is funded by the Respiratory Syncytial Virus Consortium in Europe (RESCEU), with data collection supported by the National Institute for Health Research. Pia Hardelid has attended meetings (unpaid) with MSD and Pfizer. None of the other authors have any competing interests to declare.

## Notes

### Author Declarations

This analysis was undertaken on aggregate routinely-collected data obtained from Public Health Scotland via information request and released in accordance with PHS disclosure control procedures. More information on how PHS collects and processes health data is available via their privacy notice (https://www.publichealthscotland.scot/our-privacy-notice/organisational-background/)

## References

1. Hammitt LL, Dagan R, Yuan Y, et al. Nirsevimab for Prevention of RSV in Healthy Late-Preterm and Term Infants. N Engl J Med 2022;386:837–846.

2. UK Health Security Agency. Immunisation against infectious disease - The Green Book. Available at https://www.gov.uk/government/collections/immunisation-against-infectious-disease-the-green-book#the-green-book. Accessed 17 January 2023.

3. Loyal J, Shapiro ED. Refusal of Intramuscular Vitamin K by Parents of Newborns: A Review. Hosp Pediatr 2020;10:286.

4. Golding J, Paterson M, Kinlen LJ. Factors associated with childhood cancer in a national cohort study. Br J Cancer 1990;62:304–308.

5. Klebanoff MA, Read JS, Mills JL, Shiono PH. The Risk of Childhood Cancer after Neonatal Exposure to Vitamin K. https://doi.org/101056/NEJM199309233291301 1993;329:p905-908.

6. Fear NT, Roman E, Ansell P, Simpson J, Day N, Eden OB. Vitamin K and childhood cancer: a report from the United Kingdom Childhood Cancer Study. Br J Cancer 2003;89:1228.

7. American Academy of Pediatrics Committee on Fetus and Newborn. Controversies concerning vitamin K and the newborn. American Academy of Pediatrics Committee on Fetus and Newborn - PubMed. Pediatrics 2003;112:191–192.

8. Bernhardt H, Barker D, Reith DM, Broadbent RS, Jackson PM, Wheeler BJ. Declining newborn intramuscular vitamin K prophylaxis predicts subsequent immunisation refusal: A retrospective cohort study. J Paediatr Child Health 2015;51:889–894.

9. Loyal J, Taylor JA, Phillipi CA, et al. Factors Associated With Refusal of Intramuscular Vitamin K in Normal Newborns. Pediatrics 2018;142.

10. Marcewicz LH, Clayton J, Maenner M, et al. Parental Refusal of Vitamin K and Neonatal Preventive Services: A Need for Surveillance. Matern Child Health J 2017;21:1079–1084.

11. Public Health Scotland. Child health pre-school review coverage. Available at https://www.publichealthscotland.scot/publications/child-health-pre-school-review-coverage/child-health-pre-school-review-coverage-2020-to-2021/.

12. Fraser E. Scottish Index of Multiple Deprivation 2020 - gov.scot. Available at https://www.gov.scot/collections/scottish-index-of-multiple-deprivation-2020/. Accessed 8 February 2023.

13. R Core Team, Team RDC, R Core Team. R: A Language and Environment for Statistical Computing. Vienna, Austria, Austria: R Foundation for Statistical Computing; 2017.

14. Public Health Scotland. Organisational background - Our privacy notice - Public Health Scotland. Available at https://www.publichealthscotland.scot/our-privacy-notice/organisational-background/. Accessed 16 February 2023.

15. Sahni V, Lai FY, MacDonald SE. Neonatal vitamin K refusal and nonimmunization. Pediatrics 2014;134:497–503.

16. Bora K. Gaps in the coverage of vitamin K1 prophylaxis among newborns in India: insights from secondary analysis of data from the Health Management Information System. Public Health Nutr 2021;24:5589–5597.

17. Kc A, Singh DR, Upadhyaya MK, Budhathoki SS, Gurung A, Målqvist M. Quality of Care for Maternal and Newborn Health in Health Facilities in Nepal. Matern Child Health J 2020;24:31.

18. Marcewicz LH, Clayton J, Maenner M, et al. Parental Refusal of Vitamin K and Neonatal Preventive Services: A Need for Surveillance. Matern Child Health J 2017;21:1079–1084.

19. McQuaid F, Mulholland R, Sangpang Rai Y, et al. Uptake of infant and preschool immunisations in Scotland and England during the COVID-19 pandemic: An observational study of routinely collected data. PLOS Med 2022;19:e1003916.

