## Supplementary File 1 for "Uptake of intramuscular vitamin K administration after birth and maternal and infant demographic variables: a national cohort study"

**MEDICAL-IN-CONFIDENCE**

**Health Visitor First Visit Report**

**HEALTH VISITOR COPY**

DATE OF VISIT:

NAME ADDRESS

CHI NO GENDER BIRTH WEIGHT

PLACE OF BIRTH MOTHERS D.O.B. HV NO.

HB HD TIME

.........................

.........................

.........................

.........................

...............

POSTCODE

*****

EDD

PLEASE PRINT CLEARLY IN BALL POINT PEN

**GP / TC NAME:**

**CHSP TC No.**

**SIRS TC No.**

Primary carer

Additional

carer

Current LAC Status

Bilingual/multilingual (Y/N)

Carer present with child (Y)

Other

Ethnicity:

Is English 1st language at home? (Y/N)

Schedule for Immunisation (Y/ N)

Vitamin K given

I/M (Y/N)

Oral (Y/N)

Primary carer current smoker (Y/N)

Child exposed to 2nd hand smoke (Y/N)

Concerns raised by carer, enter (R)

Feeding/Diet

Growth/Weight

Sleep

Development

Physical Health

Other

**Summary**: list any **issues** likely to be relevant to the child's ongoing health, development or well-being.

**ENTER ISSUE STATUS**

Issue Status

Read Code

PLEASE PRINT CLEARLY

(1)

(2)

(3)

(4)

**Health Plan Indicator (HPI)**

Summary comment

Practitioners involved in review (enter Y for all that apply)

HV

Staff Nurse

Nursery Nurse/FSW

GP

Other

Place of review

(enter Y for all that apply)

Home

Clinic

GP Practice

Other

Signature

Print Name

ID No

ver 1.9 230217 (ver 1.8.2 200217)

**Future action**: enter code if applicable	**P** - Provide	**S** - Signposted to	**D** - Discuss with	**R** - Request assistance from	**W** - Refused

GP	Parenting	Audiology	Speech &	Community	CAMHS	Childsmile Support	Language	Paediatrics

Smoking	Child	Early	Financial	Social Work	Physio/OT	Other Cessation	Healthy Weight	Education	Advice Services	Services

**FEEDING:-**

Ever breast fed (Y/N)	Always exclusively breast fed (Y/N)

Current feeding	(previous 24 hours)	Child's age when breast feeding stopped:	Weeks	Days (B, F, M, O, U)

TB Risk Status - Please indicate which countries the following people were born in:

Parent/Carers	(1)	(2)	*****

Grandparents	(1)	(2)	***** (where	applicable)	(3)	(4)	*****

Has BCG been given? (Y/N)	*****	If Yes, Date	*****

*Please check the information above and if appropriate, enter amendments below. Please also advise the GP of any changes.*

Change of Surname to:	Change of Forename to:

Change of Address to:	Postcode: CASELOAD HV

**MEDICAL-IN-CONFIDENCE**

**Health Visitor First Visit Report**

**COMPUTER COPY**

DATE OF VISIT:

NAME ADDRESS

CHI NO GENDER BIRTH WEIGHT

PLACE OF BIRTH MOTHERS D.O.B. HV NO.

HB HD TIME

.........................

.........................

.........................

.........................

...............

POSTCODE

*****

EDD

PLEASE PRINT CLEARLY IN BALL POINT PEN

**GP / TC NAME:**

**CHSP TC No.**

**SIRS TC No.**

Primary carer

Additional

carer

Current LAC Status

Bilingual/multilingual (Y/N)

Carer present with child (Y)

Other

Ethnicity:

Is English 1st language at home? (Y/N)

Schedule for Immunisation (Y/ N)

Vitamin K given

I/M (Y/N)

Oral (Y/N)

Primary carer current smoker (Y/N)

Child exposed to 2nd hand smoke (Y/N)

Concerns raised by carer, enter (R)

Feeding/Diet

Growth/Weight

Sleep

Development

Physical Health

Other

**Summary**: list any **issues** likely to be relevant to the child's ongoing health, development or well-being.

**ENTER ISSUE STATUS**

Issue Status

Read Code

PLEASE PRINT CLEARLY

(1)

(2)

(3)

(4)

**Health Plan Indicator (HPI)**

Summary comment

Practitioners involved in review (enter Y for all that apply)

HV

Staff Nurse

Nursery Nurse/FSW

GP

Other

Place of review

(enter Y for all that apply)

Home

Clinic

GP Practice

Other

Signature

Print Name

ID No

ver 1.9 230217 (ver 1.8.2 200217)

**Future action**: enter code if applicable	**P** - Provide	**S** - Signposted to	**D** - Discuss with	**R** - Request assistance from	**W** - Refused

GP	Parenting	Audiology	Speech &	Community	CAMHS	Childsmile Support	Language	Paediatrics

Smoking	Child	Early	Financial	Social Work	Physio/OT	Other Cessation	Healthy Weight	Education	Advice Services	Services

**FEEDING:-**

Ever breast fed (Y/N)	Always exclusively breast fed (Y/N)

Current feeding	(previous 24 hours)	Child's age when breast feeding stopped:	Weeks	Days (B, F, M, O, U)

TB Risk Status - Please indicate which countries the following people were born in:

Parent/Carers	(1)	(2)	*****

Grandparents	(1)	(2)	***** (where	applicable)	(3)	(4)	*****

Has BCG been given? (Y/N)	*****	If Yes, Date	*****

*Please check the information above and if appropriate, enter amendments below. Please also advise the GP of any changes.*

Change of Surname to:	Change of Forename to:

Change of Address to:	Postcode: CASELOAD HV

**MEDICAL-IN-CONFIDENCE**

**Health Visitor First Visit Report**

**PARENT COPY**

DATE OF VISIT:

NAME ADDRESS

CHI NO GENDER BIRTH WEIGHT

PLACE OF BIRTH MOTHERS D.O.B. HV NO.

HB HD TIME

.........................

.........................

.........................

.........................

...............

POSTCODE

*****

EDD

PLEASE PRINT CLEARLY IN BALL POINT PEN

**GP / TC NAME:**

**CHSP TC No.**

**SIRS TC No.**

Primary carer

Additional

carer

Current LAC Status

Bilingual/multilingual (Y/N)

Carer present with child (Y)

Other

Ethnicity:

Is English 1st language at home? (Y/N)

Schedule for Immunisation (Y/ N)

Vitamin K given

I/M (Y/N)

Oral (Y/N)

Primary carer current smoker (Y/N)

Child exposed to 2nd hand smoke (Y/N)

Concerns raised by carer, enter (R)

Feeding/Diet

Growth/Weight

Sleep

Development

Physical Health

Other

**Summary**: list any **issues** likely to be relevant to the child's ongoing health, development or well-being.

**ENTER ISSUE STATUS**

Issue Status

Read Code

PLEASE PRINT CLEARLY

(1)

(2)

(3)

(4)

**Health Plan Indicator (HPI)**

Summary comment

Practitioners involved in review (enter Y for all that apply)

HV

Staff Nurse

Nursery Nurse/FSW

GP

Other

Place of review

(enter Y for all that apply)

Home

Clinic

GP Practice

Other

Signature

Print Name

ID No

ver 1.9 230217 (ver 1.8.2 200217)

**Future action**: enter code if applicable	**P** - Provide	**S** - Signposted to	**D** - Discuss with	**R** - Request assistance from	**W** - Refused

GP	Parenting	Audiology	Speech &	Community	CAMHS	Childsmile Support	Language	Paediatrics

Smoking	Child	Early	Financial	Social Work	Physio/OT	Other Cessation	Healthy Weight	Education	Advice Services	Services

**FEEDING:-**

Ever breast fed (Y/N)	Always exclusively breast fed (Y/N)

Current feeding	(previous 24 hours)	Child's age when breast feeding stopped:	Weeks	Days (B, F, M, O, U)

TB Risk Status - Please indicate which countries the following people were born in:

Parent/Carers	(1)	(2)	*****

Grandparents	(1)	(2)	***** (where	applicable)	(3)	(4)	*****

Has BCG been given? (Y/N)	*****	If Yes, Date	*****

*Please check the information above and if appropriate, enter amendments below. Please also advise the GP of any changes.*

Change of Surname to:	Change of Forename to:

Change of Address to:	Postcode: CASELOAD HV
